# Striatal representations and network correlates of digital motor scores in Parkinson’s disease

**DOI:** 10.64898/2026.09.27.26364116

**Authors:** Benjamin Roeben, Dominik Blum, Barbara Gallardo Montes, Tudor M. Ionescu, Clint Hansen, Isabel Wurster, Anna-Katharina von Thaler, Kristina Herfert, Benjamin Bender, Gerhard Eschweiler, Thomas Gasser, Kathrin Brockmann, Daniela Berg, Walter Maetzler, Christian la Fougère, Matthias Reimold

## Abstract

Digital health technology (DHT) enables to detect and quantify charateristics motor dysfunction in Parkinson’s disease (PD). However, the understanding of the relationship between dopaminergic nigrostriatal denervation and quantitative composite digital motor scores (DMS) captured by DHT remains largely limited to date.

In this hybrid PET-MRI study, we used simultaneously acquired PET with ^11^C-d-threo-methylphenidate (^11^C-dMP-PET) – a tracer of presynaptic dopamine transporters – and resting-state-fMRI (rs-fMRI) to investigate this relationship and to explore associated neural network correlates in PD.

Seventeen PD patients and 39 healthy controls underwent ^11^C-dMP-PET and rs-fMRI. Quantitative composite DMS for arm swing, gait and balance were calculated using DHT-derived parameters. Voxel-based correlation analysis of ^11^C-dMP-PET binding potential (BP_NP_) was performed for each DMS and resulting clusters were used for seed-based functional connectivity analyses.

We found that DMS of arm swing, gait and balance exhibit differential striatal representations associated with dopaminergic denervation as assessed by ^11^C-dMP-PET. Specifically, lower BP_NP_ correlated with higher (i. e. more pathologic) arm swing DMS in the central putamen and higher balance DMS in the right caudate, whereas the most robust correlation was observed for lower BP_ND_ with higher gait DMS in the right caudate and anteromedial putamen. Seed-based functional connectivity analysis revealed lower functional connectivity in a frontostriatal network encompassing the basal ganglia and medial frontal regions associated to pathological gait DMS in PD.

Taken together, our study demonstrates striatal representations of *key domains of PD motor dysfunction* captured by DHT. Highlighting the complex interplay of gait motor control, we show that gait dysfunction is associated with dopaminergic denervation in the anteromedial *executive* subregion of the striatum and frontostriatal connectivity changes in regions involved in planning and initiation of movement sequences. Therefore, our study provides first preliminary insights into the relationship of dopaminergic degeneration and DMS encouraging further investigation in larger cohorts and longitudinal studies to construct and validate digital motor biomarkers obtained from DHT as outcome measures in clinical trials in PD.

**Abbreviated summary:** Using ^11^C-d-threo-methylphenidate-PET and resting-state-fMRI, Roeben et al. investigate the relationship between dopaminergic denervation and digital motor scores (DMS) in Parkinson’s disease. They show that gait DMS are associated with dopaminergic denervation in the anteromedial *executive* striatum and frontostriatal connectivity changes in regions involved in planning and initiation of movement sequences.

## Introduction

It has been increasingly recognized that subtle motor deficits in Parkinson’s disease (PD) already emerge before clinical symptoms are evident^1–4^ that would allow diagnosis of PD based on established clinical diagnostic criteria.^5^ Filling this diagnostic gap, digital health technologies (DHT) using body-worn sensor systems have been increasingly utilised in recent years to investigate motor dysfunction in PD.^6,7^ A growing number of studies using DHT have investigated specific aspects of motor dysfunction in PD. As a result, DHT-derived biomarkers have been shown to sensitively capture objective and quantitative information of various motor characteristics (e.g. arm swing, gait and balance) providing promising potential for detection and delineation of motor dysfunction in PD,^8–11^ already in early disease stages and potentially even years before the clinical diagnosis of PD.^12–21^ Consequently, detection of motor dysfunction using DHT is of highest interest, in particular validating specific DHT-derived parameters as digital motor biomarkers or composite scores of motor characteristics that might serve as novel outcomes in clinical trials.^7,22^ However, validation of outcome measures derived from DHT is still warranted. Importantly, to achieve validation of such digital motor biomarkers or composite scores of PD motor dyfunction derived from DHT-based motor assessments for clinical trials, it is necessary to verify whether distinct motor deficits captured by DHT are specifically related to dopaminergic nigrostriatal denervation in PD.

To date, the understanding of the relationship between motor characteristics derived from DHT and dopaminergic nigrostriatal denervation in terms of anatomical representations in the striatum and neural network correlates is still limited and only a few studies have investigated this relationship so far. Using ^123^I-FP-CIT dopamine transporter single-photon emission computed tomography (DAT-SPECT) and DHT-based gait assessment, Hirata et al. showed a negative correlation of dopamine transporters with to impairment of gait automaticity in the bilateral anteromedial striatum – representing the *executive* subregion of the striatum – in drug-naïve PwPD.^23^ In this study, ^123^I-FP-CIT uptake correlated with the coefficient of variation of stride length under fast walking and dual task conditions.^23^ Furthermore, two more recent studies also investigated the relationship of dopaminergic denervation with clinical and quantitative motor data. Using DAT-SPECT data, the first study showed that the onset of motor symptoms occurred first in the upper limb in a cohort of recently diagnosed PwPD.^18^ Extending these findings, the second study indicated a somatotopic pattern of dopaminergic denervation with the region with the highest degree of denervation (i.e. the caudal-intermediate subregion of the putamen) coinciding with functional representations of the upper limb using spatial covariance analysis of ^18^F-DOPA-PET and kinematic assessments of hand and foot bradykinesia in a nested subcohort of early PwPD from the aforementioned study.^24^

Beyond this and given the clinical variability of PD motor symptoms, evidence from recent studies suggests that there i s also variability of spatial and temporal DHT-derived parameters. While specific temporal parameters (e.g. step time) seem to evolve early but remain relatively stable over the course of the disease,^25^ other temporal parameters (e.g. step time variability) only occur in early to moderate stages of PD.^13,26^ Furthermore, spatial parameters (e.g. step length) rather seem to change and vary with PD progression.^41^ Therefore, in order to investigate the pathophysiological relationship of changes in both temporal and spatial motor parameters with dopaminergic denervation, it encourages to examine this relationship in PwPD at the transition from early to mid-stage PD.

In this study, we took advantage of DHT capturing quantitative data of a comprehensive set of temporal and spatial motor characteristics of *key domains of PD motor dysfunction* (i.e. arm swing, balance and gait) in a cohort of early- to mid-stage PwPD to (i) investigate representations of digital motor scores (DMS) and their association with dopaminergic denervation in the striatum using PET imaging with ^11^C-d-threo-methylphenidate (dMP-PET) – a tracer of presynaptic dopamine transporters^27,28^ – and (ii) to explore related functional connectivity changes as part of a hybrid PET-fMRI study.

## Methods

### Study design and participants

We enrolled 17 PwPD diagnosed according to the Movement Disorder Society (MDS) consensus criteria prospectively recruited from a previous MRI study at our centre (Training-PD study^29^). For the present study, 40 HC without a diagnosis of neurodegenerative disease were prospectively recruited out of the TREND study (*Tübingen Evaluation of Risk Factors for Early Detection of Neurodegeneration*).^30^ Two participants of the TREND study were diagnosed with PD in the course of the TREND study before being recruited for the present study and were thus included in the PD cohort. In short, the TREND study is a prospective longitudinal study initiated in 2009 with biennial assessments of 1201 elderly participants aged between 50 and 80 years without neurodegenerative diseases. Exclusion criteria were a diagnosis of a neurodegenerative disease, stroke, or inflammatory central nervous disease or the administration of dopaminergic or antipsychotic drugs at study inclusion. The study was performed at the Department of Neurology and the Department of Psychiatry and Psychotherapy of the University Hospital Tübingen, Germany, comprising a large comprehensive assessment battery with mainly quantitative, unobtrusive measurements. For more details about the TREND study see https://www.trend-studie.de/. Study data are collected and managed using REDCap electronic data capture tools hosted at University of Tübingen ^31^.

The study was performed in accordance with the ethical standards laid down in the 1964 Declaration of Helsinki and its later amendments. Ethical approval of the study was granted by the ethical committee of the University of Tübingen (#415/2014BO1) and written informed consent from all participants was obtained prior to study inclusion.

### Clinical assessments

Clinical assessments of all participants were performed by a neurologist specialized in movement disorders including a standardized neurological examination as well as rating of severity of motor symptoms using the motor part of the Movement Disorders Society-sponsored revision of the Unified PD Rating Scale (MDS-UPDRS II I)^32^ and the Hoehn and Yahr stage (H&Y) scale.^33^ Laterality of PD symptoms was determined based on the MDS-UPDRS III score and handedness was surveyed from the participants.

Global cognitive function was assessed with the Montréal Cognitive Assessment (MoCA).^34^ The Trail Making Test (TMT) part A was used to assess executive function and part B to evaluate psychomotor speed.^35^ Total levodopa-equivalent daily dose (LEDD) was calculated according to established conversion factors.^36,37^

### Digital motor assessments

Parameters of arm swing, balance and gait characteristics were captured using the Mobility Lab® system (OPAL APDM, Inc., Portland, OR, United States) on the same day of the PET-fMRI acquisition. The Mobility Lab® system has been validated in older adults and PwPD^9,14^ and includes six body-worn sensors: one at each wrist and ankle, one chest and one lower back sensor. Temporal and spatial parameters of range of motion, symmetry and turning measures were collected under two motor tasks and prespecified conditions:

i. *Gait:* Participants were asked to walk in a straight line over a 20 m walkway under 4 different conditions:

a. at the participants’ preferred speed
b. at fast walking speed
c. at fast walking speed while checking boxes on a sheet of paper as fast as possible (dual-tasking condition 1) and
d. dual task at fast walking speed with serial subtractions from 100 in steps of seven (dual-tasking condition 2).
ii. *Arm swing:* Parameters for arm swing were derived from gait tasks *(a)*, *(b)* and *(d)*.
iii. *Balance:* Participants were instructed to stand on a foam mat with their feet close together for 30 seconds each:

a. with eyes open and
b. with eyes closed.

The raw data of derived parameters of the respective motor characteristics (9 gait parameters per gait task, total n of gait parameters = 36; 13 balance parameters per balance task, total n of balance parameters = 26; 16 arm swing parameters from gait tasks *(a)*, *(b)* and *(d)*, total n of arm swing parameters n= 48) was stored on the Mobility Lab® system, subsequently extracted and processed using validated algorithms.^16,21,38^

### PET-MRI acquisition

After intravenous bolus injection of a maximum of 500 MBq (max. 6,1 μg) ^11^C-d-threo-methylphenidate (^11^C-dMP), simultaneous PET-MRI was performed on a 3-Tesla Biograph mMR scanner (Siemens Healthineers Erlangen, Germany). Dynamic PET images (acquisition time=60 minutes, 12 frames×10 seconds, 12 frames×20 seconds, 12 frames×60 seconds, 14 frames×180 seconds) were reconstructed using ordinary Poisson OSEM (3 iterations, 21 subsets) with ultrashort echo time-based attenuation correction and a 4 mm Gaussian filter stored in a 256×256×127 voxel grid with a voxel size of 1.4×1.4×2.0 mm³. High-resolution T1-weighted MR images were acquired using a 3D MPRAGE sequence (repetition time (TR)/inversion time (TI)/echo time (TE)=2300/900/3.37üms, flip angle=9°, voxel size=1.0×1.0×1.0ümm^3^, slices=192, field of view (FOV)=256×256ümm^2^). Resting-state fMRI (rs-fMRI) data was obtained using a gradient-echo echo-planar imaging (EPI) sequence (TR/TE=2000/30.0üms, voxel size=3.0×3.0×3.2ümm^3^, slices=36, gap=0.8 mm, volumes=300, FOV=192×192ümm^2^).

### PET image processing and kinetic modelling

PET images were preprocessed using the Statistical Parametrical Mapping software package (SPM12; Wellcome Department of Imaging Neuroscience, UCL, London, UK; https://www.fil.ion.ucl.ac.uk/spm/software/spm12), the Computational Anatomy Toolbox (CAT 12.7 (1)) and MATLAB 2020a (MathWorks, Natick, MA). A PET sum image (frames 5 min p.i. to 8 min p.i.) was used as reference to determine realignment parameters for all frames after 5 min p. i. Next, the sum image was coregistered to the MR image and transformation parameters were applied to all PET frames (i.e. to early native frames and to realigned frames). Using CAT, MR images were then normalised to MNI space using DARTEL to the IXI555 MNI template with 161×197×161 voxels of 1 mm cubic size. Deformation maps derived from the DARTEL normalisation was applied to the PET data without modulation. Normalized PET images were smoothed with an 8 mm Gaussian kernel. ^11^C-dMP binding potential (BP_NP_) was calculated using custom in-house MATLAB scripts applying the multilinear reference tissue model 2 with occipital cortex as reference region and washout k2’=0.05 min^−1^. For review of a schematic depiction of the processing pipeline see Supplementary Figure 1.

### Preprocessing of resting-state fMRI

Preprocessing of rs-fMRI data was performed using the SPM-based CONN toolbox (version 22a; RRID:SCR_009550).^39^ Co-registered structural and fMRI data were processed following the default preprocessing pipeline implemented in the CONN toolbox including functional realignment, unwarping, slice-timing correction, outlier identification, direct segmentation, normalization and functional smoothing.^40^

### Statistical analysis

Statistical analysis of demographic and clinical data was performed using SPSS statistical software version 28.0 (IBM Corp., Armonk, NY). Data distribution was assessed using Shapiro-Wilk tests, Q-Q plots and histograms. Group comparisons were performed using Student’ s t-test for parametric continuous data and Pearson χ^2^ test for categorical data. Intergroup comparisons of clinical scores were calculated using univariate analysis of variance with covariates age and sex. All statistical tests were two-sided and *p* values ≤ 0.05 were considered statistically significant.

#### Calculation ofcomposite Digital Motor Scores

To select parameters to be used for the calculation of the composite DMS for arm swing right, arm swing left, gait and balance, we performed two-sample t-tests for all parameters comparing the PD and HC group. Parameters that showed at least moderate effect sizes (Cohen’s d>0.5; for arm swing in at least one side, i.e. left or right) were selected for calculation of the composite DMS. The DMS was calculated by summing all selected parameters. Before summing, each parameter was z-transformed to ensure that the DMS was not dominated by parameters with high variance and for each parameter, the sign was chosen so that higher values reflect more pathological scores. Parameter values exceeding two standard deviations (SD; SD calculated from the PD group) were considered outliers and replaced by “NaN” (not a number). If one “NaN” appeared in the set of parameters used to calculate the DMS, it was replaced by its group mean before linear combination. If more than two parameters were “NaN”, the respective DMS was also replaced by “NaN”.

#### Correlations with dMP-PET binding potential

We performed correlation analyses of BP_ND_ with disease duration, MDS-UPDRS III, MoCA and TMT scores to assess the relationship of each clinical marker with dopaminergic denervation.

To localize striatal representations associated with DMS, we calculated voxel-wise multiple regressions for each DMS in the PD group using SPM (voxel-level threshold p < 0.01, cluster size >10 voxels). The goal was to detect functionally specific regions, while trivial results are conceivable due to correlations between DMS and disease stage on the one side and between disease stage and BP_ND_ in striatal subregions on the other, with resulting coordinates reflecting regions with low variance rather than functional specificity. Our approach to this challenge was twofold: first, we calculated the BP_ND_ in the whole striatum as an index for disease stage and included this index as a covariate in the statistical model (covariates: age, whole striatum BP_ND_). Second, we used the SPM toolbox MASCOI to obtain masked contrast images from T-maps (here: contrast = regression coefficient for the respective DMS) using the concept of a regionally least significant contrast^41^ and report the coordinates of the maximum contrast instead of the maximum T-value. Using BP_ND_ as a nuisance variable also has the effect of reducing variance in terms of global factors of dopamine transporter expression related neither to neurodegeneration nor to motor function.

For further analysis, we focused on the gait DMS as – when accounting for whole striatum BP_ND_ – this was the only DMS for which we observed a “negative” correlation (i.e. reduced BP_ND_ associated with more pathological DMS) and where our results were robust insofar as we also observed a significant effect at the same coordinates without including whole striatum BP_ND_. To illustrate the relation between BP_ND_ and gait at the coordinate calculated with SPM and the MASCOI toolbox, irrespective of whole striatum BP_ND_, we plotted age-corrected BP_ND_ over gait DMS for the HC and PD groups. For age correction, we used a linear model and corrected individual BP_ND_ to reflect the corresponding BP_ND_ at the age of 66.8 years (mean age in HC and Parkinson’s disease). To avoid a post-hoc bias, in this analysis, we refrained from reporting *p*-values in PwPD, however, we did test for a correlation in the HC group. Furthermore, for exploration, we repeated the SPM analysis of gait DMS correlates by using the left-right flipped image volumes of patients with clinically left-dominant manifestation.

#### Seed-to-voxel functional connectivity analysis

In order to examine functional connectivity changes related to the gait DMS, the gait-associated cluster resulting from the dMP-PET analysis was selected for a seed-to-voxel analysis using the CONN toolbox. We used the coordinates of the gait DMS associated PET cluster (MNI coordinates [x/y/z]: 16/15/-1) as a seed to investigate (i) functional connectivity in the PD and HC group separately in a first step and (ii) the effect of the gait DMS on functional connectivity to all other brain voxels in a second level analysis.

### Ethical Standards

The study was performed in accordance with the ethical standards laid down in the 1964 Declaration of Helsinki and its later amendments. Ethical approval of the study was granted by the ethical committee of the University of Tübingen (#415/2014BO1) and written informed consent from all participants was obtained prior to study inclusion.

### Data availability

The data of this study are available upon request to the corresponding author.

## Results

### Demographic and clinical characteristics

Seventeen PwPD and 39 HC were included in this study. Demographic and clinical characteristics of study participants are shown in Table 1. The PD group was significantly younger (62.1±7.5 years vs. 68.9±5.8 years; *p*<0.001) and had significantly fewer females (23.5% vs. 56.4%; *p*=0.023) than the HC group. Mean disease duration in the PD group was 8.3±3.0 years. MDS-UPDRS III and LEDD were significantly higher in the PD group (both *p*<0.001) as expected. There were no significant differences in handedness, MoCA score as well as in TMT part A and B scores and delta TMT B-A.

### Digital Motor Scores

The discriminative analysis of temporal and spatial parameters obtained from the DHT-based motor assessment comparing the PD and HC groups yielded 6 balance parameters, 3 gait parameters as well as 13 parameters for arm swing right and 13 parameters for arm swing left (Table 2; see Supplementary Table 1 for a detailed overview of all DHT parameters). Balance encompassed temporal and spatial parameters from both conditions with eyes closed as well as with eyes open. For gait and arm swing, the results of the analysis comprised temporal and spatial parameters under normal walking as well as fast walking and fast walking with dual tasking (serial subtractions). The resulting parameters were used to calculate DMS for arm swing, gait and balance (see Table 2).

In the PD group, the DMS for right and left arm swing showed a significant positive correlation (*r*=0.79), while the gait DMS was negatively correlated with right arm swing DMS (*r*=-0.82) and left arm swing DMS (*r*=-0.56). The Balance DMS was neither correlated with the gait DMS nor with the DMS for right and left arm swing.

### 11C-dMP-PET and correlations with clinical and Digital Motor Scores

In PD, dMP-PET showed the typical pattern of a pronounced reduction of BP_ND_ in the posterior parts of the striatum. Furthermore, there was no overlap of the striatum BP_ND_ between the PD and HC group (PD: 0.72 [range: 0.51-1.00]) vs HC: 1.65 [range: 1.20-1.93]). The median asymmetry index was 3% (range: 0 – 11%). Disease duration was negatively correlated with BP_ND_ with the strongest effect in the right caudate (*r*=-0.55; *p*=0.021). MDS-UPDRS III was negatively correlated with BP_ND_ with the strongest effect in the left putamen (*r*=-0.51; *p*=0.035), consistent with the higher frequency of predominantly right-sided clinical manifestation in the PD group (see Table 1). There were no significant correlations for MoCA, TMT A, TMT B and delta TMT B-A (see Supplementary Table 2).

The correlation analysis of BP_ND_ with the DMS for arm swing, gait and balance showed differential anatomical representations for each DMS in the striatum (see figure panel in Table 3). First, higher (i. e. more pathologic) gait DMS was significantly correlated with lower dMP-PET BP_ND_ in a cluster in the right anteromedial putamen and ventral caudate nucleus (*Tmax*=-3.07; p=0.005). Here, we found a negative correlation of the gait DMS and BP_ND_ (r=0.54), while in HC the gait DMS showed a positive correlation with BP_ND_ (*r*=0.43, p=0.010; Figure 1). When using the flipped images, we observed a virtually identical cluster with a slightly higher *T_max_* of −3.35 (p=0.003).

**Figure 1:**
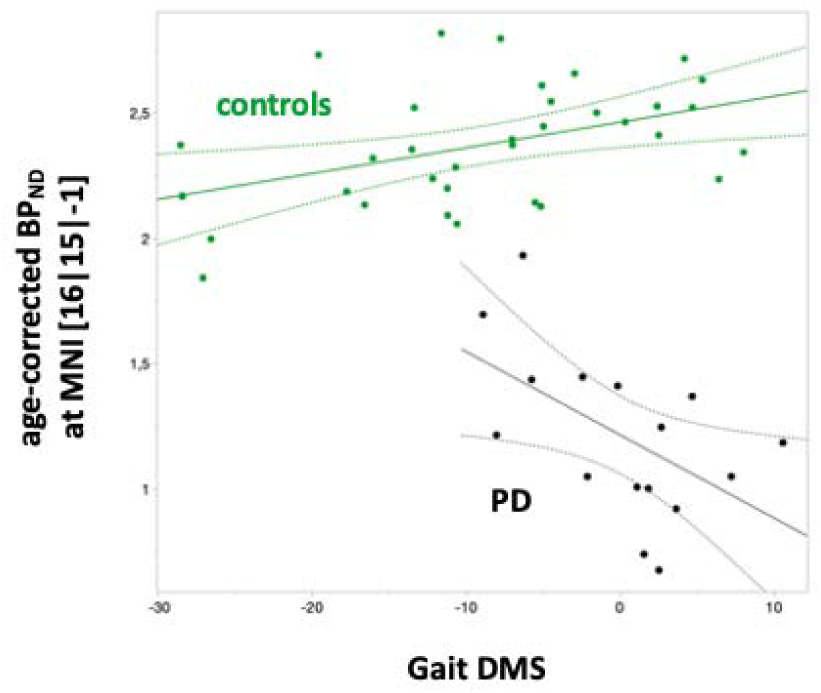
Correlation of dMP-PET binding potential (BP_ND_) with the gait DMS. In PD, lower dMP-PET age-corrected BP_ND_ correlated with higher (i.e. more pathologic) gait DMS (r=-0.54) at the coordinate of the maximum effect (maximum beta coefficient; SPM analysis and MASCOI toolbox, see table 2). Interestingly, in HC, we observed an inverse correlation (*r*=0.43, p=0.010). We refrain from reporting a *p*-value for the correlation effect in the PD group since the coordinate was derived from the SPM analysis.

In contrast to the findings for the gait DMS, lower BP_ND_ at the same location was associated with lower DMS of arm swing right (*T*max=4.16; *p*=0.001) and left (*T*max=3.67; *p*=0.002), while generally, in the remaining striatum, lower BP_ND_ was associated with higher arm swing DMS. Specifically, the strongest correlation for arm swing were observed for the DMS of right arm swing and the contralateral central putamen (*T*max=-6.10; *p*<0.001). Finally, higher, more pathologic balance DMS was associated with lower BP_ND_ in a small cluster in the right caudate nucleus (*T*max=-3.10; *p*=0.005).

### Seed-based functional connectivity analysis

Since the gait DMS related cluster showed the most robust effect in the analysis of the dMP-PET data, we used its coordinate (i.e. the coordinate of the maximum correlation coefficient within the cluster) as a seed in a seed-to-voxel functional connectivity analysis and assessed the effect of the gait DMS on functional connectivity in a second level analysis.

As a first step, we investigated the functional connectivity between the coordinates of the gait DMS associated PET cluster as a seed and all other brain voxels in the PD and HC groups separately. This showed negative correlations with sensorimotor cortex and positive correlations in a network encompassing the basal ganglia and medial frontal regions in both the PD and HC group (see Supplementary figure 2).

The second level analysis, assessing the effect of gait DMS on functional connectivity, revealed *negative* correlations in the bilateral basal ganglia and medial frontal regions (Figure 2), i. e. in regions where first level analysis showed positive correlations. These correlations were observed in PD only. Specifically, more pathological gait in PD was associated with weaker functional connectivity in this network encompassing the putamen, pallidum, caudate and nucleus accumbens as well as medial frontal regions (i.e. orbitofrontal cortex, medial frontal cortex (MFC) and the Anterior Cingulate Cortex (ACC) including the Cingular Motor Areas (CMA); for detailed data see Supplementary Table 3).

**Figure 2:**
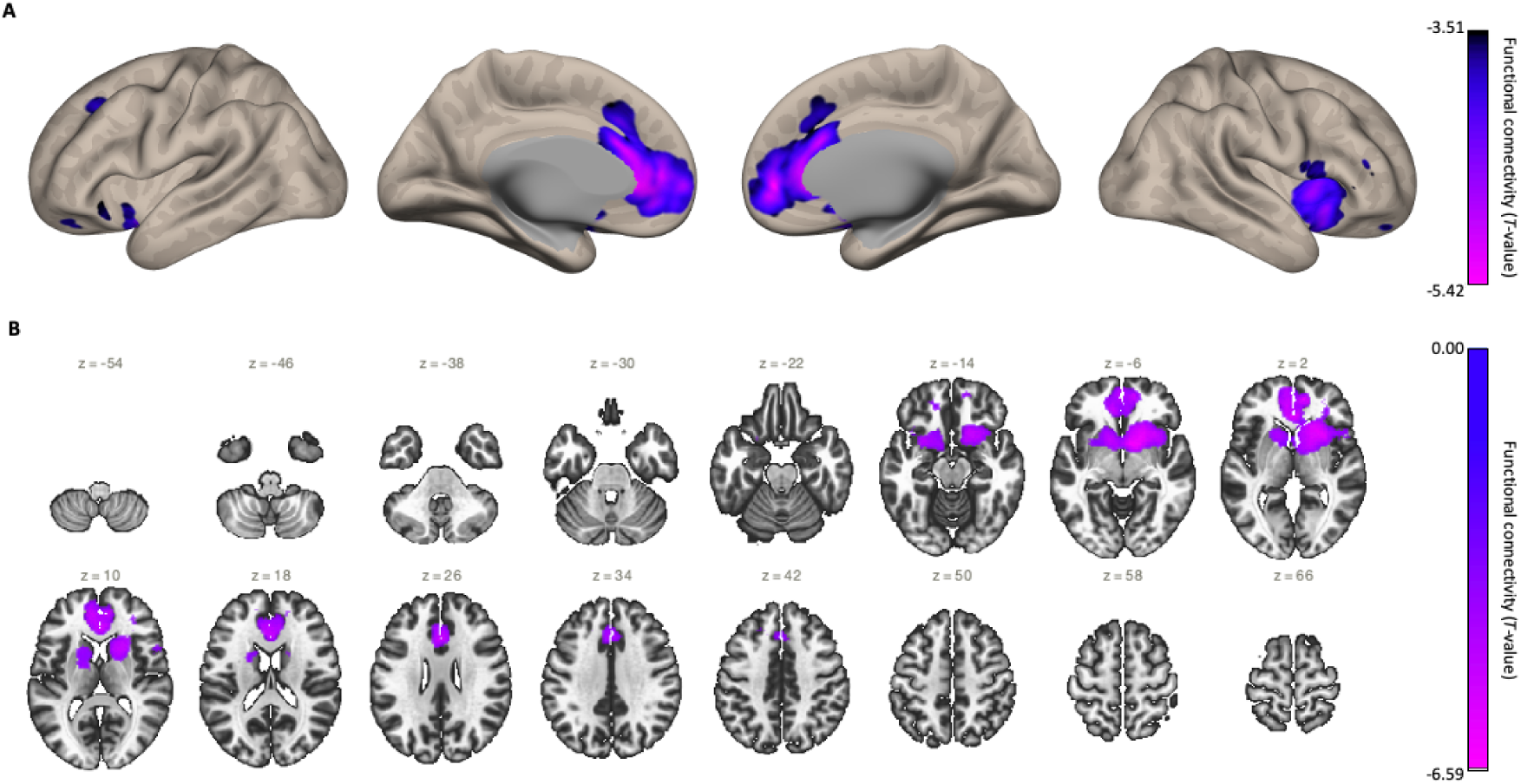
Seed-based functional connectivity analysis. Using the coordinates of the gait DMS associated PET cluster (MNI coordinates [x/y/z]: 16/15/-1) as a seed, the effect of the gait DMS on functional connectivity to all other brain voxels was investigated (voxel threshold *p*<0.001 (uncorrected) and false discovery rate (FDR) corrected cluster threshold *p*<0.05). PD patients showed weaker functional connectivity in a network encompassing the putamen, pallidum, caudate and nucleus accumbens (see multi-slice view panel; B) as well as medial frontal cortical regions (see inflated cortical surface view; A).

## Discussion

In this hybrid PET-MRI study, we used simultaneously acquired ^11^C-dMP-PET and rs-fMRI to investigate the relationship between quantitative composite DMS of *multiple key domains of PD motor dysfunction* captured by DHT – namely arm swing, gait and balance – with nigrostriatal dopaminergic denervation, and to explore associated neural network correlates in PD.

Our study shows that (i) DMS of arm swing, gait and balance exhibit differential striatal representations associated with dopaminergic denervation as assessed by ^11^C-dMP-PET; (ii) pathologic gait DMS are robustly correlated with dopaminergic denervation in the right ventral caudate nucleus and the adjacent anteromedial putamen in PD; and (iii) pathological gait DMS in PD is related to lower functional connectivity in a network encompassing the basal ganglia and medial frontal regions.

So far, previous studies using DHT have predominantly analysed correlations of clinical measures or functional anatomy of striatal subregions with respect to PD motor dysfunction only using single DHT-derived motor parameters.^13–15,17,19,23,25,42^ However, it is still unclear which DHT-derived motor parameters best reflect neuronal dysfunction in a subregion-specific way. Our approach of using composite DMS as a basis for image analysis offers some advantages, while the chosen methodology to select and combine single motor score requires some justification. Given the high number of single motor parameters (total n=110) recorded in this study, it is beyond the scope of an usual PET study to rigorously evaluate possible combinations of motor scores using a purely data driven approach. In the present study, we therefore grouped DHT-derived motor parameters into a priory defined categories (arm swing, gait and balance) and selected motor parameters using an univariate approach. The latter is crucial, since multivariate approaches, including principal component analysis (PCA), common e.g. for the development of a diagnostic marker in early stages, would introduce a considerable risk of overfitting. Parameters that showed at least moderate effect sizes (Cohen’s d>0.5 in two-sample t-test PD versus HC) and were thus chosen for calculation of the composite DMS in our study included parameters that have been shown to delineate and track PD-related motor symptoms in previous DHT studies (e.g. temporal and spatial DHT-derived motor parameters reflecting velocity/pace, variability and asymmetry).^13,16,17^

As a result, our analysis revealed differential anatomical representations of DMS of arm swing, gait and balance associated with dopaminergic denervation in the striatum. Specifically, higher (i.e. more pathologic) arm swing DMS of the left side was associated with lower BP_ND_ in ipsilateral and contralateral putaminal clusters. Higher arm swing DMS on the right side was correlated with lower BP_ND_ in a contralateral cluster in the centromedial putamen and a contralateral cluster in the caudate nucleus, whereas higher balance DMS was associated with lower BP_ND_ in a small cluster in the right caudate nucleus.

The most robust finding of our study is represented by the significant negative correlation of the gait DMS with the BP_ND_ in the right caudate and anteromedial putamen in PwPD, that is, higher (i.e. more pathologic) gait DMS correlated with lower BP_ND_ in the PD group as expected. Conversely, in HC, we found a *positive* correlation of gait DMS with BP_ND_ in the same striatal subregion. This can be interpreted as a confirmation that this subregion is indeed involved in regulation of gait motor processes. Consequently, one hypothesis explaining the positive correlation in HC could be upregulation of dopamine transporters due to reduced dopaminergic tone in different conditions negatively impacting gait motor control. Furthermore, given the strong correlation between BP_ND_ in this region and the gait DMS, the finding that, in PD, BP_ND_ in this region was *positively* correlated with DMS of arm swing might also indicate compensatory changes of arm swing in the context of impaired gait.^43–45^

Our finding of a negative correlation of BP_ND_ with the gait DMS and the location of the maximum effect are well in line with findings of another recent study relating the caudate and anteromedial putamen to impairment of gait automaticity.^23^ Using ^123^I-FP-CIT DAT-SPECT and DHT-based gait assessment, Hirata et al. showed a negative correlation of dopamine transporters with the coefficient of variation of stride length in the bilateral anteromedial striatum – representing the *executive* subregion of the striatum – in drug-naïve PwPD.^23^

The voxel-wise analyses of both our and the study by Hirata et al. revealed correlations of BP_ND_ in the anteromedial striatum. While our study showed a right-sided cluster, the study by Hirata et al. showed a predominantly left-sided pattern. This may be explained by the higher proportion of patients with clinically left predominant PD symptoms in the study by Hirata et al., in contrast to the higher proportion of PwPD with clinically right predominant PD symptoms in our study. In line with this, left-right flipping of PET images of patients with clinically left predominant PD symptoms further increased the observed correlation of dMP-PET BP_ND_ and the gait DMS in our study. In other words, the correlation was highest between the gait DMS and the less affected striatum.

The less robust correlations of BP_ND_ with the DMS of arm swing and balance, might be due to different reasons. The balance DMS was neither correlated with the gait DMS nor with the DMS for right and left arm swing. However, while the clinical manifestation of postural instability typically occurs in more advanced disease stages, subtle disturbances of balance and postural control might already be detected in early stages of PD.^9^ Pointing to this, our analysis showed a negative correlation with BP_ND_ in a small cluster in the caudal part of the posteromedial putamen – the subregion of the putamen showing earliest disease-related neurodegeneration.^46–48^

Furthermore, other non-dopaminergic neurotransmitter systems might also have additional contributions to the pathophysiology of PD-related symptoms, e.g. the noradrenergic, ^49,50^ serotonergic^51^ and cholinergic systems^52–57^. For instance, there is mounting evidence for an association of cholinergic degeneration with gait impairment ^52–58^. Consequently, dysfunction of non-dopaminergic neurotransmitter systems might also have relevant influence on other PD motor symptoms including alterations of arm swing and balance. ^59^

Using the gait associated DMS cluster in the right anteromedial striatum as a seed, we found positive correlations with a frontostriatal network in both the PD and the HC groups. In contrast, this network showed weaker correlations associated with more pathological gait assessing the effect of the gait DMS on functional connectivity in the PD group. Specifically, this network encompassed the putamen, pallidum, caudate, accumbens as well as insular, orbitofrontal and medial frontal cortex (MFC) including frontal premotor regions (i.e. the pre-Supplementary Motor Area (pre-SMA) and the Cingular Motor Areas (CMA) of the Anterior Cingulate Cortex (ACC)).

In agreement with the subregional organization of the striatum, the anterior subregion of the putamen together with the caudate is described to represent the *executive* part of the putamen exhibiting strong connections to frontal regions.^60–62^ Distinct gait-related metabolic networks including frontal regions have been described based on spatial covariance analysis of fluoro-deoxy-glucose (FDG) PET and quantitative gait characteristics. ^63^ Previous studies investigating the relationship of striatal dopamine depletion with functional connectivity have shown functional connectivity changes of a network including the striatum, mesolimbic cortex and sensorimotor regions related to PD symptoms.^62,64,65^ Furthermore, specific profiles of functional connectivity of the anterior putamen with the pre-SMA and ACC as well as the caudate nucleus with the dorsal prefrontal cortex (dPFC) have been shown by comparing the connectivity of the anterior compared to the posterior putamen.^62,65^

In addition, it is well established that frontostriatal circuits are centrally involved in motor planning and initiation as well as motivation processing,^66,67^ which is particularly relevant in PD.^68^ Specifically, planning and initiation of movement sequences have been related to the MFC,^69–72^ the pre-SMA^73,74^ and the ACC.^75^ Importantly, these neocortical regions are the first to show Lewy body pathology and related neurodegeneration,^76^ and lesions of these regions have been described to cause akinetic symptoms^77^ as well as interference with control over voluntary action and sequences of movements.^73,74^ In line with this, studies investigating cortico-striatal connectivity using fMRI found reduced functional connectivity of these regions associated to the clinical manifestation of akinesia^78^ and response to dopaminergic treatment in PD.^79,80^ Furthermore, deep brain stimulation has been shown to improve bradykinesia and rigidity^81^ and notably stride time recorded using DHT^82^ by targeted stimulation of tracts connected to supplementary motor and premotor cortices.

Finally, the ACC has been described as a central hub for the integration of motor and cognitive information as well as aspects of drive and motivation.^83^ Connections of the ACC and pre-SMA with other premotor and prefrontal regions (e.g. dorsal premotor cortex (dPMC) and dorsolateral prefrontal cortex (DLPFC)) have been associated with executive function and cognitive motor control in terms of action selection and planning.^84,85^ While executive dysfunction has been shown to be an early and characteristic feature of cognitive deterioration in PD,^86,87^ this i s also particularly relevant in terms of gait motor control as executive dysfunction has been shown to contribute to impairment of gait performance,^88,89^ in particular under dual-tasking conditions.^90,91^ Highlighting this complex interplay of gait and cognition, a recent study showed that the association of dual-task gait and cognitive impairment is mediated by smaller grey matter volume in the anterior and middle cingulate cortices.^92^ Furthermore, a study on patients with cerebral small vessel disease demonstrated strong interrelations of cognitive impairment, apathy and gait dysfunction associated with damage to meso-cortical pathways as common underlying pathophysiology,^93^ which could be also relevant in PD.

Taken together, connectivity of frontostriatal circuits have been shown to play a pivotal role in the pathophysiology of gait dysfunction in PD due to several reasons:

i. frontal premotor regions (e.g. MFC, pre-SMA and ACC including the Cingulate Motor Areas (CMA)) are centrally involved in the planning and initiation of movement sequences as well as in self-paced movements, which might be particularly relevant in terms of gait impairment in PD (e.g. FOG);
ii. executive dysfunction i s a predominant feature of early cognitive deterioration in PD mediated by frontal regions (e.g. ACC and DLPFC) contributing to gait dysfunction, e. g. under dual tasking conditions; and
iii. as the ACC has been implemented in internally cued, extended effortful behaviours, it might also be involved in motivational aspects for movement initiation in PD.^94^

In summary, the findings of our combined dMP-PET-fMRI analysis highlight the relationship of dopaminergic denervation and frontostriatal connectivity changes related to gait dysfunction, supporting mounting evidence that suggest a network-dependent progression of PD symptoms.^95–97^ Our findings are further supported by recent evidence indicating that variability of PD motor symptoms is influenced by (compensatory) cortical mechanisms of premotor cortical networks rather than basal ganglia dysfunction alone.^44^ Finally, this might also have relevant clinical implications for targeting specific PD symptoms with personalized both invasive and non-invasive neuromodulation of frontal premotor regions and/or frontostriatal circuits (e.g. with DBS).^81,82,98,99^

We acknowledge some limitations of our study. First, the number of PwPD enrolled in our study is relatively small diminishing the statistical power and generalisability of our findings. Nonetheless, given the extent of our study protocol and the effort involved for the patients, in particular with regard to the PET-MRI examination and the sensor-based assessment, patient numbers of our PD group are comparable to those of similar PET-MRI studies. Therefore, we argue that our study provides first preliminary insights into the relationship of DMS with dopaminergic degeneration and associated functional connectivity changes encouraging further investigations. Future studies with larger numbers of PwPD could confirm our findings and might also include data driven approaches beyond the scope of the present manuscript.

Second, PwPD were assessed in medication ON state of their regular dopaminergic medication. In turn, potential effects of the dopaminergic medication on functional connectivity might cover disease-related effects in our rs-fMRI analysis. However, sustained medication effects might even persist after short term withdrawal of the dopaminergic medication, while longer periods in the OFF state may not be tolerated by PwPD due to symptom worsening, making it impossible for patients to participate in such an elaborate study. Furthermore, the study results might also be altered due to excessive fatigue in the OFF state. It is conceivable that assessment in dopaminergic OFF state could yield even more pronounced effects in line with our findings.

However, while the findings of our rs-fMRI analysis are largely consistent with the results of fMRI studies with drug-naïve PwPD^65^ and investigations of changes in functional connectivity in the ON state compared to the OFF state,^79,80^ inference on disease-related functional connectivity changes in the OFF state is beyond the scope of this study. As PD patients in real-world scenarios and also in already ongoing clinical trials with potentially disease-modifying treatments (e.g. the PASADENA clinical trial; https://clinicaltrials.gov/study/NCT03100149) are typically ON dopaminergic medication, we argue that i t is of equally high relevance and importance to investigate functional connectivity in PD patients ON medication to identify alterations of functional connectivity associated with disease progression that might also serve as biomarkers of treatment response in clinical trials.

Finally, the binding of the dMP tracer is not expected to be altered by the dopaminergic medication.^100^

Third, the DHT-based motor assessment was conducted before the PET-fMRI. Therefore, we cannot infer direct conclusions on the relationship of motor sequences with functional connectivity due to the comprehensive protocol of the DHT-based assessment “offline” as opposed to motor task-related fMRI protocols. However, by first investigating correlations of DMS and dMP-PET BP_ND_, and subsequent seed-based functional connectivity analysis of resulting clusters, we argue that this approach allowed us to gain insight into the relationship between the constructed DMS and dopaminergic denervation as well as associated functional connectivity changes.

The strengths of our study include the standardized and comprehensive DHT-based motor assessment providing objective and quantitative data of different motor characteristics. Finally, the PET-fMRI protocol was simultaneously acquired on a hybrid 3-Tesla PET-MRI scanner including a high-resolution T1-weighted sequence enabling optimal spatial registration and a time-efficient examination providing enhanced patient comfort.

Taken together, our study demonstrates differential anatomical representations of *multiple key domains of PD motor dysfunction* captured by DHT in the striatum. Highlighting the complex interplay of gait motor control, we conclude that gait dysfunction is associated with dopaminergic denervation in the anteromedial *executive* subregion of the striatum and frontostriatal connectivity changes in regions involved in planning and initiation of movement sequences.

Digital motor biomarkers or composite scores show great potential to enable delineation of specific PD motor symptoms (e.g. gait) and therefore might provide novel, valuable outcome parameters for clinical trials. Our study provides first preliminary insights into the relationship of dopaminergic degeneration and DMS encouraging further investigation in larger cohorts and longitudinal studies to construct and validate digital motor biomarkers obtained from DHT as potential novel outcome measures in clinical trials in PD.

## Data Availability

The data of this study are available upon request to the corresponding author.

## Acknowledgements

We thank the participants for their continued participation in the TREND study and for providing biosamples. We acknowledge the work of numerous (doctoral) students and study nurses, who actively contributed to study organization, data collection, entry and monitoring.

The TREND study is being conducted at Tübingen University Hospital and has been or is supported by the Hertie Institute for Clinical Brain Research, the German Center for Neurodegenerative Diseases, the Geriatric Center of Tübingen, the Center for Integrative Neuroscience, Teva Pharmaceutical Industries, Union Chimique Belge, Janssen Pharmaceuticals, and the International Parkinson Fund. The supporting institutions had no influence on the design, conduct, or analysis of this study.

## Funding

This study was supported by the AKF (Applied Clinical Research) programme of the Faculty of Medicine of the University of Tübingen (grant F.1353098).

## Author contributions

B.R. contributed to conceptualizsation, project administration, investigation, methodology, data curation, formal analysis, writing-original draft, writing-review & editing and visualization. D.Bl., B. G-M., T.M.I and C.H. contributed to methodology, formal analysis, writing-review & editing. I.W. contributed to investigation, writing-review & editing. A-KvT was responsible for project administration, data curation, writing-review & editing. K.H. and B.B. contributed to methodology, supervision, writing-review & editing. T. G. contributed to supervision, writing-review & editing. K. B., G. E. and D.Be. contributed to resources, funding acquisition, writing-review & editing. W.M. and C.l.F. were responsible for conceptualization, funding acquisition, resources, supervision, writing-review & editing. MR was responsible for conceptualization, supervision, investigation, methodology, data curation, formal analysis, writing-original draft, writing-review & editing and visualization.

## Financial Disclosures

Dr. Roeben was supported by the Clinician Scientist program of the Medical Faculty of the University of Tübingen (grant #478-0-0).

Dr. Blum, Mrs. Gallardo-Montes, Dr. Ionescu, Dr. Hansen and Dr. von Thaler have nothing to disclose.

Dr. Wurster received funding from the Michael J. Fox Foundation in form of the Edmond J. Safra Fellowship in Movement Disorders.

Prof. Dr. Herfert has received or receives financial support as part of a collaborative study with the MODAG GmBH, unrelated to this study.

Prof. Dr. Bender has no conflicts of interest related to this study; unrelated to this work he serves as CTO and share-holder of AIRAmed GmbH.

Prof. Dr. Eschweiler has received research funding German Innovationsfonds (Fund of the Federal Joint Committee, Gemeinsamer Bundesausschuss, G-BA Grants no. VF1_2016-201).

Prof. Dr. Gasser serves on the editorial board of the Journal of Parkinson’s disease. He holds a patent re: KASPP (LRRK2) Gene, its Production and Use for the Detection and Treatment of Neurodegenerative Diseases. Prof. Gasser has received speaker’s honoraria from UCB Pharma, Novartis, Sanofi, and MedUpdate. He has received consulting fees from Bayer AG, BlueRock Therapeutics, and Biogen. He is Chairman of the Scientific Advisory Board of the “Joint Programming for Neurodegenerative Diseases” program, funded by the European Commission. He has received grant support from the German Research Foundation (DFG), the German Federl Ministry of Education and Research (BMBF), the Ministry of Science, Research and Art Baden-Württemberg (MWK), the European Commissino, the Helmholtz Association, and the Michael J. Fox Foundation.

Prof. Dr. Brockmann has received or receives funding from the Michael J. Fox Foundation for Parkinson’s Research, the German Society of Parkinson’s disease (DPG), the Health Forum Baden Wuerttemberg, the Else Kröner Fresenius Foundation, the University of Tuebingen, and from the German Research Foundation (DFG). She serves as a consultant for F. Hoffman-La Roche Ltd., Vanqua Bio, and the Michael J. Fox Foundation for Parkinson’s Research and has received speaker honoraria form Abbvie, Lundbeck, UCB and Zambon.

Prof. Dr. Berg has received or receives research grants from the German Federal Ministry of Education and Research (BMBF), the German Research Council (DFG), the Else Kröner Fresenius foundation, the Jan von Appen Foundation, the Michael J. Fox Foundation (MJFF), UCB Pharma GmbH and the European Union, all not related to this study.

Prof. Dr. Maetzler receives or received funding from the European Union, the German Federal Ministry of Education and Research, German Research Council, Michael J. Fox Foundation, Robert Bosch Foundation, Neuroalliance, Lundbeck, Sivantos and Janssen. He received speaker honoraria from Abbvie, Bayer, BIAL, GlaxoSmithKline, Heel, Licher MT, Rölke Pharma, Takeda and UCB, was invited to Advisory Boards / Consultancies of Abbvie, Aptar Digital Health, Atheneum, BIAL, Biogen, Kyowa Kirin, Lundbeck and Pfizer. He serves as an advisory board member of the Critical Path for Parkinson’ s Consortium, and an editorial board member of Geriatric Care. He serves as the co-chair of the MDS Technology Working Group.

Prof. Dr. la Fougère reports honorary speaker invitations by AAA, ImaginAB, Novartis, Recordati and Siemens Healthineers, Consultancy arrangements by ADACP, Astra Zeneca, Bayer, Ipsen, Novartis and Telix, as well as third party fundings for the Department from Novartis, ABX and Siemens Healthineers.

Dr. Reimold has nothing to disclose.

## Competing interests

The authors report no competing interests.

## Abbreviations

PD: Parkinson’s Disease
HC: Healthy Controls
DHT: Digital Health technology
DMS: Digital Motor Score
MDS-UPDRS: Movement Disorders society sponsored revision of the Unified Parkinson’s Disease Rating Scale
MoCA: Montréal Cognitive Assessment

## Notes

### Competing Interest Statement

Dr. Roeben was supported by the Clinician Scientist program of the Medical Faculty of the University of Tuebingen (grant #478-0-0).
Dr. Blum, Mrs. Gallardo-Montes, Dr. Ionescu, Dr. Hansen and Dr. von Thaler have nothing to disclose.
Dr. Wurster received funding from the Michael J. Fox Foundation in form of the Edmond J. Safra Fellowship in Movement Disorders.
Prof. Dr. Herfert has received or receives financial support as part of a collaborative study with the MODAG GmbH, unrelated to this study.
Prof. Dr. Bender has no conflicts of interest related to this study; unrelated to this work he serves as CTO and share-holder of AIRAmed GmbH.
Prof. Dr. Eschweiler has received research funding German Innovationsfonds (Fund of the Federal Joint Committee, Gemeinsamer Bundesausschuss, G-BA Grants no. VF1_2016-201).
Prof. Dr. Gasser serves on the editorial board of the Journal of Parkinsons disease. He holds a patent re: KASPP (LRRK2) Gene, its Production and Use for the Detection and Treatment of Neurodegenerative Diseases. Prof. Gasser has received speaker honoraria from UCB Pharma, Novartis, Sanofi, and MedUpdate. He has received consulting fees from Bayer AG, BlueRock Therapeutics, and Biogen. He is Chairman of the Scientific Advisory Board of the Joint Programming for Neurodegenerative Diseases program, funded by the European Commission. He has received grant support from the German Research Foundation (DFG), the German Federal Ministry of Education and Research (BMBF), the Ministry of Science, Research and Art Baden-Wuerttemberg (MWK), the European Commissino, the Helmholtz Association, and the Michael J. Fox Foundation.
Prof. Dr. Brockmann has received or receives funding from the Michael J. Fox Foundation for Parkinsons Research, the German Society of Parkinsons disease (DPG), the Health Forum Baden Wuerttemberg, the Else Kroener Fresenius Foundation, the University of Tuebingen, and from the German Research Foundation (DFG). She serves as a consultant for F. Hoffman-La Roche Ltd., Vanqua Bio, and the Michael J. Fox Foundation for Parkinsons Research and has received speaker honoraria form Abbvie, Lundbeck, UCB and Zambon.
Prof. Dr. Berg has received or receives research grants from the German Federal Ministry of Education and Research (BMBF), the German Research Council (DFG), the Else Kroener Fresenius foundation, the Jan von Appen Foundation, the Michael J. Fox Foundation (MJFF), UCB Pharma GmbH and the European Union, all not related to this study.
Prof. Dr. Maetzler receives or received funding from the European Union, the German Federal Ministry of Education and Research, German Research Council, Michael J. Fox Foundation, Robert Bosch Foundation, Neuroalliance, Lundbeck, Sivantos and Janssen. He received speaker honoraria from Abbvie, Bayer, BIAL, GlaxoSmithKline, Heel, Licher MT, Roelke Pharma, Takeda and UCB, was invited to Advisory Boards / Consultancies of Abbvie, Aptar Digital Health, Atheneum, BIAL, Biogen, Kyowa Kirin, Lundbeck and Pfizer. He serves as an advisory board member of the Critical Path for Parkinson's Consortium, and an editorial board member of Geriatric Care. He serves as the co-chair of the MDS Technology Working Group.
Prof. Dr. la Fougere reports honorary speaker invitations by AAA, ImaginAB, Novartis, Recordati and Siemens Healthineers, Consultancy arrangements by ADACP, Astra Zeneca, Bayer, Ipsen, Novartis and Telix, as well as third party fundings for the Department from Novartis, ABX and Siemens Healthineers.
Dr. Reimold has nothing to disclose.

### Author Declarations

Ethical approval of the study was granted by the ethical committee of the University of Tuebingen (#415/2014BO1).

