## Supplementary Material for "Striatal representations and network correlates of digital motor scores in Parkinson’s disease"

### Supplementary Figure 1: Schematic depiction of the PET-fMRI processing and study analysis pipeline

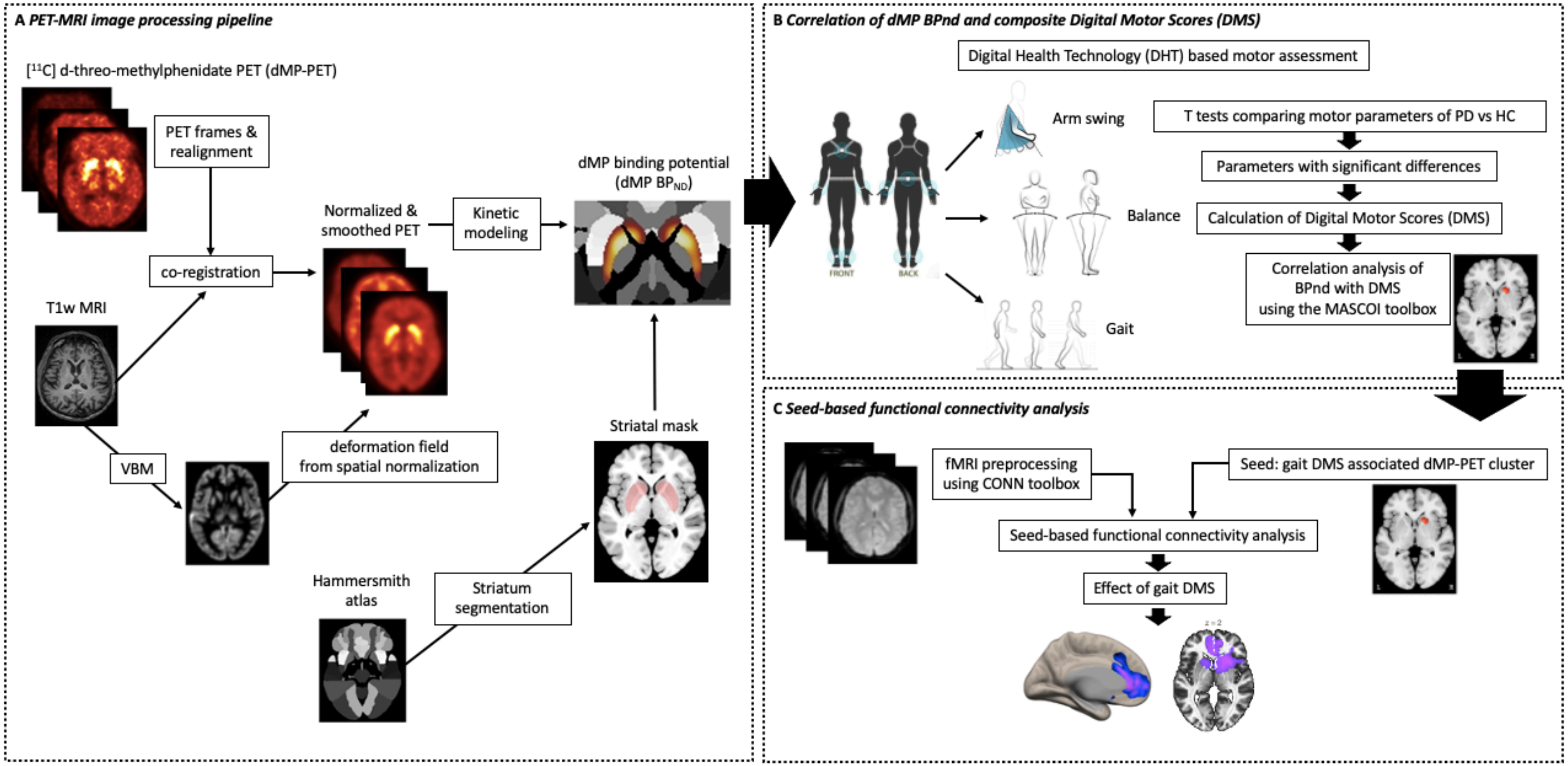


**Supplementary figure 1: Schematic depiction of the PET-fMRI processing and study analysis pipeline. (A)** A $^{11}$C-d-threo-methylphenidate (dMP) PET sum image (frames 5 min p. i. to 8 min p. i.) was used as reference to determine realignment parameters for all frames after 5 min p. i. The PET sum image was coregistered to the MR image and transformation parameters were applied to all PET frames. MR images were then normalised to MNI space using DARTEL to the IXI555 MNI template (161 × 197 × 161 voxels of 1 mm cubic size) and deformation maps derived from the DARTEL normalisation were applied to the PET data without modulation. Normalized PET images were smoothed with an 8 mm Gaussian kernel. $^{11}C$-dMP binding potential ($BP_{ND}$) was calculated applying the multilinear reference tissue model 2 with occipital cortex as reference region and washout k2' = 0.05 $min^{-1}$. **(B)** Using parameters with significant differences comparing the PD and HC group composite digital motor score (DMS) were calculated for arm swing right, arm swing left, gait and balance. Correlations of the respective DMS with $BP_{ND}$ were performed using the SPM toolbox MASCOI. **(C)** Coordinates of the gait DMS associated PET cluster (MNI coordinates [x/y/z]: 16/15/-1) were used as a seed to investigate (i) functional connectivity in the PD and HC group separately and (ii) the effect of the gait DMS on functional connectivity to all other brain voxels in a second level analysis.

**Supplementary Figure 2: Seed-based functional connectivity first level analysis in PD and HC**

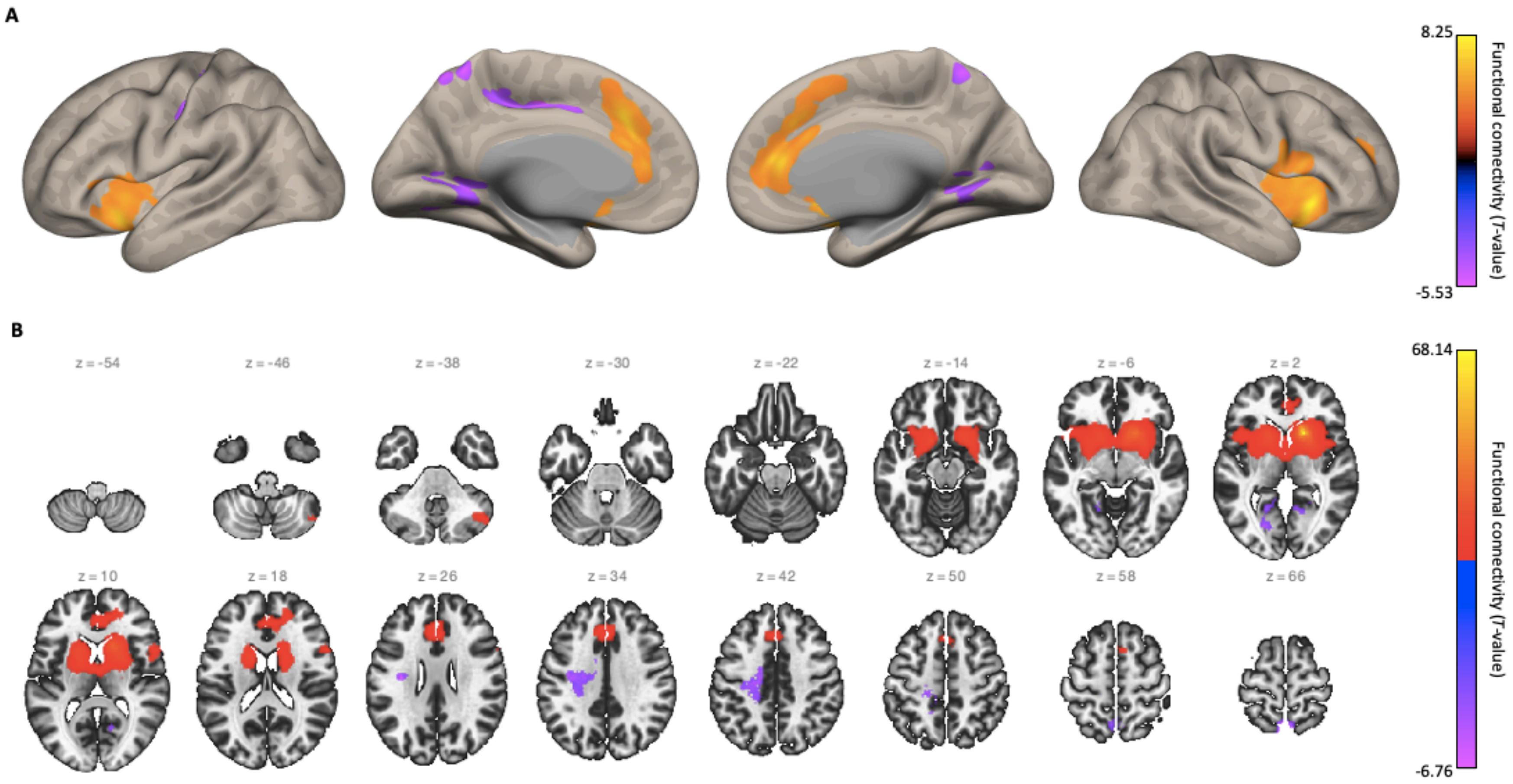

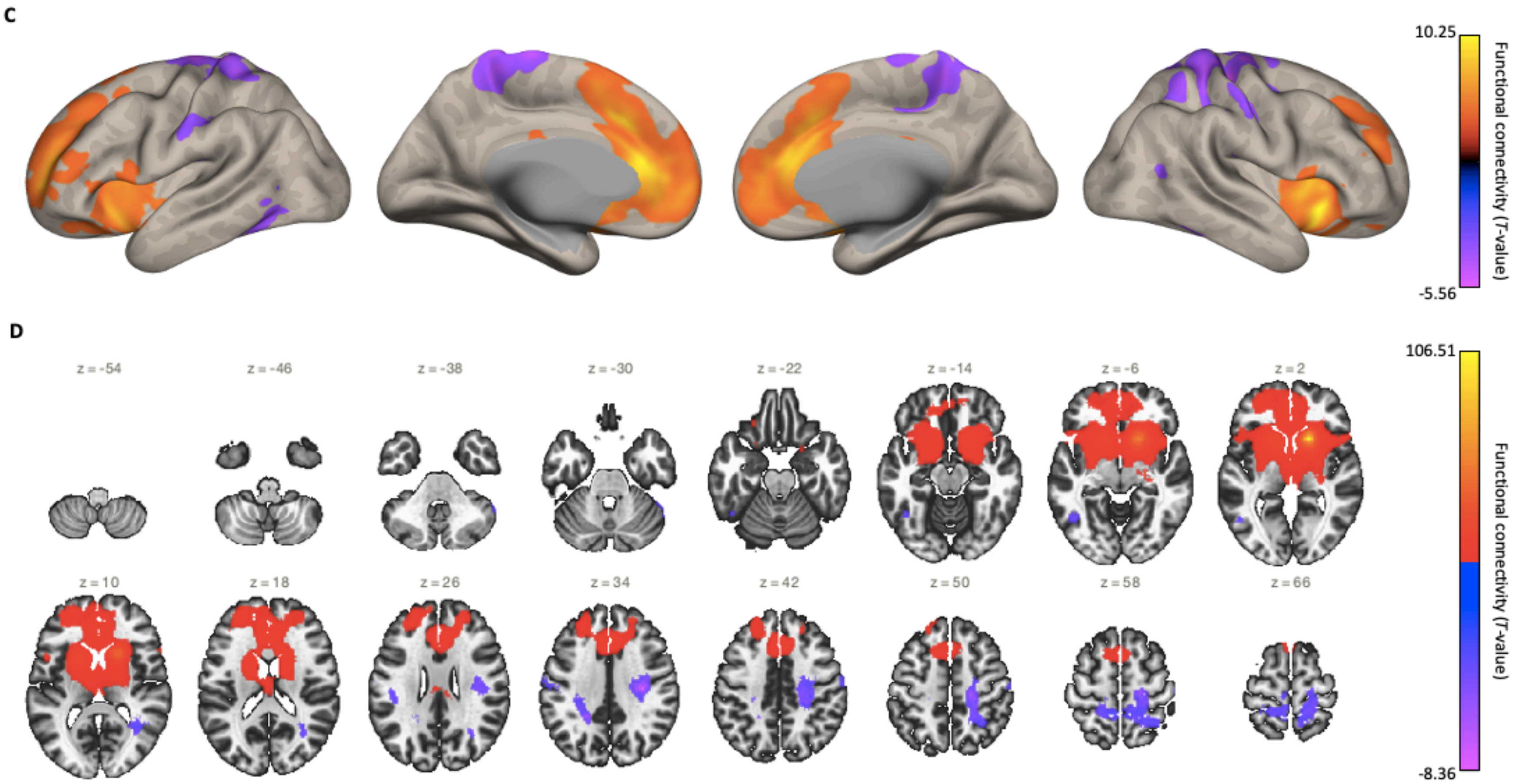


**Supplementary figure 2: Seed-based functional connectivity first level analysis in PD and HC.** First level analysis seed-to-voxel analysis investigating functional connectivity between the coordinates of the gait DMS associated PET cluster (MNI coordinates [x/y/z]: 16/15/-1) as a seed and all other brain voxels (voxel threshold $p<0.001$ (uncorrected) and false discovery rate (FDR) corrected cluster threshold $p<0.05$). Both in the PD and the HC group, negative correlations with sensorimotor cortex (see inflated cortical surface view; PD: A; HC: C) and positive correlations in a network encompassing the basal ganglia (see multi-slice view panel; PD: B; HC: D) and medial frontal cortical regions (PD: A; HC: C) were observed.

**Supplementary Table 1: Overview of parameters from the Digital Health Technology (DHT) based motor assessment and comparison between PD patients and healthy controls.**

| | **PD (n= 17)** | **HC (n= 39)** | ***Cohen's d*** |
|---|---|---|---|
| **BALANCE** | | | |
| ***Eyes open*** | | | |
| Jerk [$m^2/s^5$] | 3.92 (0.98) | 0.98 (1.96) | -0.435 |
| Mean Distance from COP trajectory [$cm/s^2$] | 11.77 (10.79) | 9.81 (3.92) | -0.367 |
| Root mean square of COP time series [$cm/s^2$] | 15.69 (14.71) | 11.77 (4.9) | -0.454 |
| Sway total path length of COP trajectory [$m/s^2$] | 24.51 (28.92) | 17.65 (10.10) | -0.383 |
| Mean velocity COP [cm/s] | 107.87(127.49) | 78.45 (39.23) | -0.383 |
| Jerk anteroposterior [$cm^2/s^3$] | 0.49 (0.1) | 0.1 (0.1) | ***-0.988*** |
| Acceleration anteroposterior [$cm/s^2$] | 9.81(10.79) | 6.86 (2.94) | -0.438 |
| Velocity anteroposterior [cm/s] | 28.44 (27.46) | 18.63 (14.71) | **-0.534** |
| Jerk mediolateral [$cm^2/s^3$] | 0.49 (0.69) | 0.1 (0.1) | ***-0.934*** |
| Acceleration mediolateral [$cm/s^2$] | 10.79 (10.79) | 7.85 (3.92) | -0.390 |
| Velocity mediolateral [cm/s] | 36.28 (23.54) | 32.36 (26.48) | -0.157 |
| Sway area from COP per time unit [$cm^2/s^5$] | 3.92 (9.81) | 0.98 (0.98) | -0.441 |
| Mean frequency [Hz] | 1.42 (0.90) | 1.31 (0.70) | -0.135 |
| ***Eyes closed*** | | | |
| Jerk [$m^2/s^5$] | 12.75 (18.63) | 10.79 (17.65) | -0.152 |
| Mean Distance from COP trajectory [$cm/s^2$] | 25.5 (14.71) | 22.56 (9.81) | -0.278 |

| | | | |
|---|---|---|---|
| Root mean square of COP time series [cm/s$^2$] | 31.38 (20.59) | 25.5 (10.79) | -0.397 |
| Sway total path length of COP trajectory [m/s$^2$] | 49.32 (26.77) | 53.93 (34.22) | 0.146 |
| Mean velocity COP [cm/s] | 215.75 (117.68) | 235.36 (147.1) | 0.138 |
| Jerk anteroposterior [cm$^2$/s$^3$] | 1.18 (1.08) | 0.29 (0.29) | **-1.369** |
| Acceleration anteroposterior [cm/s$^2$] | 21.57 (15.69) | 17.65 (8.83) | -0.328 |
| Velocity anteroposterior [cm/s] | 56.88 (44.13) | 37.27 (28.44) | **-0.581** |
| Jerk mediolateral [cm$^2$/s$^3$] | 1.08 (0.98) | 0.29 (0.39) | **-1.183** |
| Acceleration mediolateral [cm/s$^2$] | 21.57 (16.67) | 18.63 (7.85) | -0.322 |
| Velocity mediolateral [cm/s] | 64.72 (66.69) | 55.9 (40.21) | -0.184 |
| Sway area from COP per time unit [cm$^2$/s$^5$] | 11.77 (16.67) | 6.86 (6.86) | -0.442 |
| Mean frequency [Hz] | 1.41 (0.67) | 1.62 (0.57) | 0.359 |
| **GAIT** | | | |
| ***Normal walking speed*** | | | |
| Double limb support variability [s] | 0.12 (0.05) | 0.11 (0.05) | -0.085 |
| Number of steps [n] | 116.6 (8.1) | 116.1 (8.2) | -0.056 |
| Step Time [s] | 0.55 (0.04) | 0.55 (0.04) | -0.039 |
| Stride Time [s] | 1.10 (0.07) | 1.10 (0.09) | -0.031 |
| Stance Time [s] | 0.97 (0.06) | 0.96 (0.06) | -0.041 |
| Swing Time [s] | 0.13 (0.01) | 0.13 (0.02) | -0.033 |
| Double limb support [s] | 0.42 (0.03) | 0.42 (0.03) | -0.034 |
| Asymmetry [s] | 0.03 (0.04) | 0.02 (0.01) | **-0.587** |
| Step Time Variability [s] | 0.13 (0.05) | 0.12 (0.05) | -0.194 |
| ***Fast walking speed*** | | | |
| Double limb support variability [ms] | 0.08 (0.04) | 0.11 (0.05) | **0.650** |

| | | | |
|---|---|---|---|
| Number of steps [n] | 133.7 (12.0) | 130.6 (11.0) | -0.267 |
| Step Time [s] | 0.48 (0.04) | 0.49 (0.04) | 0.302 |
| Stride Time [s] | 0.96 (0.09) | 0.98 (0.08) | 0.281 |
| Stance Time [s] | 0.85 (0.08) | 0.87 (0.06) | 0.374 |
| Swing Time [s] | 0.11 (0.02) | 0.11 (0.03) | -0.078 |
| Double limb support [s] | 0.37 (0.04) | 0.38 (0.03) | -0.110 |
| Asymmetry [s] | 0.02 (0.01) | 0.01 (0.01) | -0.158 |
| Step Time Variability [s] | 0.08 (0.04) | 0.11 (0.05) | **0.615** |
| ***Fast walking speed + crosses*** | | | |
| Double limb support variability [s] | 0.12 (0.06) | 0.12 (0.06) | 0.047 |
| Number of steps [n] | 119.5 (12.6) | 122.1 (12.7) | 0.210 |
| Step Time [s] | 0.54 (0.06) | 0.53 (0.05) | -0.235 |
| Stride Time [s] | 1.08 (0.12) | 1.05 (0.11) | -0.232 |
| Stance Time [s] | 0.94 (0.10) | 0.93 (0.09) | -0.190 |
| Swing Time [s] | 0.13 (0.02) | 0.12 (0.02) | -0.390 |
| Double limb support [s] | 0.41 (0.03) | 0.40 (0.04) | -0.130 |
| Asymmetry [s] | 0.03 (0.03) | 0.02 (0.02) | -0.473 |
| Step Time Variability [s] | 0.13 (0.06) | 0.13 (0.06) | 0.003 |
| ***Fast walking speed + subtractions*** | | | |
| Double limb support variability [s] | 0.12 (0.05) | 0.11 (0.04) | -0.130 |
| Number of Steps [n] | 118.1 (15.2) | 120.8 (14.0) | 0.189 |
| Step Time [s] | 0.55 (0.07) | 0.53 (0.05) | -0.225 |
| Stride Time [s] | 1.09 (0.13) | 1.07 (0.11) | -0.206 |
| Stance Time [s] | 0.96 (0.11) | 0.94 (0.09) | -0.164 |

| | | | |
|---|---|---|---|
| Swing Time [s] | 0.13 (0.02) | 0.13 (0.03) | -0.320 |
| Double limb support [s] | 0.41 (0.05) | 0.41 (0.03) | -0.136 |
| Asymmetry [s] | 0.02 (0.01) | 0.02 (0.02) | 0.291 |
| Step Time Variability [s] | 0.12 (0.05) | 0.12 (0.04) | -0.095 |
| ***ARM SWING RIGHT*** | | | |
| ***Normal walking speed*** | | | |
| Frequency [Hz] | 0.96 (0.12) | 1.00 (0.14) | 0.243 |
| Amplitude [°] | 17.8 (9.1) | 27.3 (16.1) | **0.663** |
| Peak angular velocity [°/s] | 70.8 (31.1) | 106.6 (56.7) | **0.709** |
| Peak angular velocity forward [°/s] | 72.4 (35.9) | 103.9 (57.1) | **0.609** |
| Peak angular velocity backward [°/s] | 68.8 (30.3) | 109.4 (58.3) | **0.788** |
| Coordination between the arms [0-1] | 0.80 (0.11) | 0.78 (0.12) | -0.155 |
| Regularity of the angular velocity [0-1] | 0.81 (0.09) | 0.77 (0.10) | -0.352 |
| Percentage of swing time [%] | 84.8 (20.9) | 91.0 (10.1) | 0.436 |
| ***Fast walking speed*** | | | |
| Frequency [Hz] | 1.06 (0.10) | 1.09 (0.13) | 0.183 |
| Amplitude [°] | 21.3 (11.8) | 32.1 (18.3) | **0.655** |
| Peak angular velocity [°/s] | 80.0 (39.8) | 120.1 (63.8) | **0.698** |
| Peak angular velocity forward [°/s] | 84.2 (45.0) | 121.0 (65.1) | **0.618** |
| Peak angular velocity backward [°/s] | 75.9 (36.8) | 119.1 (64.4) | ***0.756*** |
| Coordination between the arms [0-1] | 0.80 (0.12) | 0.78 (0.14) | -0.143 |
| Regularity of the angular velocity [0-1] | 0.79 (0.10) | 0.77 (0.11) | -0.174 |
| Percentage of swing time [%] | 82.1 (22.8) | 87.6 (17.7) | 0.282 |
| ***Fast walking speed + subtractions*** | | | |

| | | | |
|---|---|---|---|
| Frequency [Hz] | 0.97 (0.4) | 1.00 (0.09) | 0.285 |
| Amplitude [°] | 20.0 (15.2) | 34.4 (16.5) | **0.893** |
| Peak angular velocity [°/s] | 79.4 (52.9) | 132.4 (58.1) | **0.588** |
| Peak angular velocity forward [°/s] | 87.3 (62.6) | 133.5 (60.6) | **0.752** |
| Peak angular velocity backward [°/s] | 74.5 (45.1) | 131.1 (58.0) | **1.038** |
| Coordination between the arms [0-1] | 0.79 (0.10) | 0.79 (0.10) | 0.046 |
| Regularity of the angular velocity [0-1] | 0.78 (0.12) | 0.77 (0.10) | -0.066 |
| Percentage of swing time [%] | 80.7 (22.6) | 92.4 (12.0) | **0.747** |
| ***ARM SWING LEFT*** | | | |
| ***Normal walking speed*** | | | |
| Frequency [Hz] | 1.00 (0.18) | 0.99 (0.15) | -0.049 |
| Amplitude [°] | 27.8 (17.9) | 36.9 (15.6) | **0.663** |
| Peak angular velocity [°/s] | 104.2 (61.0) | 130.6 (45.1) | **0.699** |
| Peak angular velocity forward [°/s] | 107.0 (66.6) | 128.7 (45.5) | **0.609** |
| Peak angular velocity backward [°/s] | 101.6 (56.5) | 132.6 (47.3) | **0.788** |
| Coordination between the arms [0-1] | 0.83 (0.10) | 0.85 (0.10) | 0.193 |
| Regularity of the angular velocity [0-1] | 0.82 (0.07) | 0.82 (0.10) | 0.0 |
| Percentage of swing time [%] | 89.3 (17.6) | 96.6 (5.9) | **0.677** |
| ***Fast walking speed*** | | | |
| Frequency [Hz] | 1.08 (0.11) | 1.06 (0.11) | -0.194 |
| Amplitude [°] | 32.5 (18.6) | 43.2 (16.0) | **0.635** |
| Peak angular velocity [°/s] | 118.9 (64.9) | 148.1 (45.5) | **0.556** |
| Peak angular velocity forward [°/s] | 123.4 (70.0) | 150.5 (45.3) | **0.498** |
| Peak angular velocity backward [°/s] | 114.5 (60.8) | 145.7 (48.1) | **0.596** |

| | | | |
|---|---|---|---|
| Coordination between the arms [0-1] | 0.83 (0.12) | 0.85 (0.10) | 0.138 |
| Regularity of the angular velocity [0-1] | 0.81 (0.10) | 0.82 (0.10) | 0.158 |
| Percentage of swing time [%] | 92.9 (10.5) | 96.5 (8.1) | 0.407 |
| ***Fast walking speed + subtractions*** | | | |
| Frequency [Hz] | 0.96 (0.14) | 1.00 (0.11) | 0.304 |
| Amplitude [°] | 30.9 (30.7) | 40.4 (18.5) | 0.416 |
| Peak angular velocity [°/s] | 105.0 (84.7) | 145.1 (59.9) | **0.588** |
| Peak angular velocity forward [°/s] | 108.9 (86.9) | 148.4 (64.6) | **0.548** |
| Peak angular velocity backward [°/s] | 101.0 (84.0) | 145.1 (55.9) | **0.670** |
| Coordination between the arms [0-1] | 0.82 (0.11) | 0.83 (0.11) | 0.060 |
| Regularity of the angular velocity [0-1] | 0.80 (0.10) | 0.77 (0.17) | -0.172 |
| Percentage of swing time [%] | 89.6 (16.3) | 97.1 (4.7) | **0.764** |

Values are presented as mean ± standard deviation of parameters before normalization. Parameters for the respective DMS were selected based on at least moderate effect sizes (Cohen's d > 0.5) and are presented in bold font. For arm swing, parameters with moderate effect size on at least one side were selected for arm swing DMS of both sides. HC = healthy control subjects; PD = Parkinson's disease patients; COP = Center of pressure.

### Supplementary Table 2: Correlation of dMP-PET BPnd and clinical scores in the PD group

| | **Left** | | | | **Right** | | | |
|---|---|---|---|---|---|---|---|---|
| | **Caudate** | | **Putamen** | | **Caudate** | | **Putamen** | |
| | *r* | *p* | *r* | *p* | *r* | *p* | *r* | *p* |
| Disease duration | ***-0.43*** | ***0.082*** | -0.39 | 0.119 | **-0.55** | **0.021** | ***-0.48*** | ***0.054*** |
| MDS-UPDRS III | -0.31 | 0.224 | **-0.51** | **0.035** | -0.11 | 0.673 | -0.21 | 0.408 |
| MoCA | +0.20 | - | +0.06 | - | +0.22 | - | -0.05 | 0.858 |
| TMT A | -0.05 | 0.851 | +0.10 | - | -0.23 | 0.385 | - | 0.881 |
| TMT B | -0.16 | 0.532 | -0.02 | 0.941 | -0.35 | 0.174 | -0.13 | 0.616 |
| Delta TMT B-A | -0.19 | 0.458 | -0.06 | 0.814 | -0.37 | 0.148 | -0.18 | 0.482 |

Significant results are shown in bold for one-sided $p<0.05$. Trends are presented in italicized bold font for $p<0.10$. Regions of interest for the caudate and putamen were defined based on the Hammersmith atlas without thresholding.

### Supplementary Table 3: Anatomical regions from the seed-to-voxel fMRI analysis in the PD group

| Anatomical region | Hemisphere | Voxels within the anatomical region (% of activation) | | |
|---|---|---|---|---|
| | | PD | HC | 2nd level analysis |
| | | | | PD |
| ***Subcortical / Basal ganglia*** | | | | |
| Putamen | right | 653 (81) | 743 (92) | 412 (52) |
| | left | 650 (75) | 746 (86) | 229 (26) |
| Caudate | right | 449 (86) | 455 (87) | 316 (61) |
| | left | 478 (89) | 499 (93) | 324 (60) |
| Pallidum | right | 158 (59) | 247 (92) | 104 (39) |
| | left | 143 (47) | 197 (65) | 22 (7) |
| Thalamus | right | 200 (16) | 611 (48) | 22 (2) |
| | left | 198 (15) | 616 (45) | 33 (2) |
| Accumbens | right | 84 (100) | 84 (100) | 84 (100) |
| | left | 107 (100) | 107 (100) | 106 (99) |
| Amygdala | right | 38 (11) | 137 (40) | - |
| | left | 39 (12) | 110 (34) | 8 (2) |
| ***Limbic lobe*** | | | | |
| Cingulate gyrus, anterior division | - | 799 (31) | 1462 (56) | 1341 (52) |
| Cingulate gyrus, posterior division | - | 8 (0) | 8 (0) | - |
| Paracingulate cortex | right | 385 (28) | 1113 (82) | 685 (41) |
| | left | 333 (25) | 1170 (89) | 556 (52) |

| | | | | |
|---|---|---|---|---|
| Subcallosal cortex | - | 141 (13) | 330 (29) | 124 (11) |
| ***Frontal lobe*** | | | | |
| Insular cortex | right | 393 (29) | 543 (40) | 368 (27) |
| | left | 316 (24) | 432 (32) | 41 (3) |
| Orbitofrontal cortex | right | 213 (15) | 526 (36) | 156 (11) |
| | left | 164 (10) | 594 (35) | 107 (6) |
| Medial frontal Cortex (MFC) | - | - | 239 (24) | 137 (14) |
| Frontal pole | right | 29 (0) | 663 (8) | 64 (1) |
| | left | - | 2023 (29) | 135 (2) |
| Superior frontal gyrus | right | 83 (3) | 233 (9) | - |
| | left | 49 (2) | 520 (18) | 33 (1) |
| Middle frontal gyrus | right | - | 6 (0) | - |
| | left | - | 79 (3) | 2 (0) |
| Inferior frontal gyrus, pars opercularis | right | - | 82 (12) | 78 (11) |
| | left | 53 (7) | 109 (14) | - |
| Inferior frontal gyrus, pars triangularis | right | - | 16 (3) | 8 (1) |
| | left | - | 31 (5) | - |
| Frontal opercular cortex | right | 7 (2) | 112 (36)<br>115 (32) | - |
| | left | 158 (45) | | - |
| Central opercular cortex | right | 55 (6) | 14 (2) | 1 (0) |
| | left | 23 (2) | | - |
| Precentral gyrus | right | 66 (2) | 487 (11) | 2 (0) |
| | left | 23 (1) | 415 (10) | - |

| | | | | |
|---|---|---|---|---|
| Juxtapositional lobule cortex | right | 2 (0) | 16 (2) | - |
| (Supplementary motor cortex) | left | - | 23 (4) | - |
| ***Temporal Lobe*** | | | | |
| Temporal pole | right | - | 8 (0) | - |
| | left | 8 (0) | 24 (1) | - |
| Middle temporal gyrus, temporooccipital | right | - | 6 (1) | - |
| part | left | - | 16 (2) | - |
| Inferior temporal gyrus, temporooccipital | right | - | 13 (2) | - |
| part | left | - | 70 (10) | - |
| Temporo-occipital fusiform cortex | right | - | - | - |
| | left | - | 49 (8) | - |
| Parahippocampal gyrus, anterior divison | right | - | 4 (1) | - |
| | left | - | - | - |
| ***Parietal Lobe*** | | | | |
| Postcentral gyrus | right | - | 698 (22) | - |
| | left | 6 (0) | 755 (21) | - |
| Superior parietal lobule | right | - | 424 (29) | - |
| | left | - | 34 (2) | - |
| Parietal opercular cortex | right | - | 2 (0) | - |
| | left | - | - | - |
| Precuneus | - | - | 123 (2) | - |
| ***Occipital Lobe*** | | | | |
| Lingual gyrus | right | 97 (6) | - | - |
| | left | 91 (6) | - | - |

| | | | | |
|---|---|---|---|---|
| Intracalcarine cortex | right | 26 (3) | - | - |
| | left | 31 (5) | - | - |
| Lateral occipital cortex, superior division | right | 2 (0) | 1 (0) | - |
| | left | 17 (0) | - | - |
| Lateral occipital cortex, inferior division | right | - | - | - |
| | left | - | 22 (1) | - |
| Occipital fusiforum gyrus | right | - | - | - |
| | left | 2 (0) | - | - |
| ***Cerebellum*** | | | | |
| Crus I | right | 89 (4) | 192 (8) | - |
| | left | - | - | - |
| Crus II | right | 126 (6) | 35 (2) | - |
| | left | - | - | - |
| Lobule IV/V | right | - | - | - |
| | left | 5 (1) | - | - |
| Lobule VI | right | - | 17 (1) | - |
| | left | - | - | - |
| Lobule VIIb | right | 3 (1) | - | - |
| | left | - | - | - |
